# Fisher-Rao distance and sex differences in disease prevalence trajectories

**DOI:** 10.1101/2025.11.14.25340249

**Authors:** José Manuel Rodríguez Caballero

**Affiliations:** Département de mathématiques et de statistique, Université Laval

**Keywords:** Statistics as Topic, Entropy, Sex Factors, Prevalence, Global Burden of Disease

## Abstract

**Objectives:** We introduce a new application of the Fisher–Rao geodesic distance to quantify sex differences in age-stratified chronic-disease prevalence trajectories, modelling those trajectories as dynamical systems on the hyperbolic plane and using GBD 2021 data.

**Methods:** We analysed prevalence for 10 major chronic conditions across three regions—US states (50 states + DC), 24 Western European countries, and 47 Japanese prefectures—over 1990–2019. We logit-transformed prevalence and summarised each age-sex cohort by normal-approximation parameters (*µ, σ*), which were then embedded in the hyperbolic plane. Sex differences were quantified as the difference between the total Fisher–Rao trajectory lengths for males and females. We assessed cross-regional consistency using parametric (mean differences) and nonparametric (Cohen’s g) summaries, and compared Fisher–Rao results to KL divergence, absolute mean differences, and absolute SD differences.

**Study design:** Cross-sectional analysis of GBD 2021 prevalence data modelled as trajectories in the hyperbolic plane.

**Results:** The Fisher–Rao distance showed greater cross-regional consistency than the alternative metrics. Males showed greater trajectory shifts in neoplasms, cardiovascular diseases, chronic respiratory diseases, diabetes/kidney diseases, skin/subcutaneous diseases, and sense organ diseases. Females showed greater shifts in neurological disorders, mental disorders, and substance use disorders. Digestive diseases exhibited mixed patterns.

**Conclusions:** This geometry-informed metric outperforms alternatives in assessing sex disparities in disease burdens, enhancing public health surveillance and equity in chronic disease management. Future extensions should incorporate gender dimensions.

## 1 Introduction

The Fisher–Rao metric has its roots in Fisher’s 1922 Fisher information matrix for statistical estimation. [1]. Rao (1945) formalised this idea as a metric tensor on statistical manifolds and introduced geodesic distances now known as the Rao distance. [2] This laid the groundwork for information geometry, advanced by Shun-ichi Amari in the 1980s via dual affine connections and applications to machine learning and neural networks [3].

Recent work applies the Fisher–Rao metric in biological contexts such as generative modelling and optimal transport. The FISHER-FLOW model employs Fisher-Rao reparameterisation for generative modelling over discrete data [4], evaluated on DNA promoter and enhancer sequences. Related studies optimise cosine schedules in masked diffusion models for biological sequences [5] and extend to *L*^*p*^-Fisher-Rao metrics with Amari-Čencov *α*-connections for Finsler geometries [6]. The Wasserstein-Fisher-Rao metric models optimal transport in leaf venation, blood circulation, and neuronal networks [7].

In shape analysis, a core area of statistical learning, the Fisher-Rao metric facilitates unsupervised clustering of biological objects by quantifying shape similarities [8]. In phylogenetics, it arises from character distributions under two-state symmetric Markov models on trees [9]. It also supports functional data registration for neuroscience spike trains and gene expression [10].

This paper presents a novel epidemiologic application of the Fisher-Rao geodesic distance to quantify sex-based differences in chronic disease prevalence trajectories across age groups, aligning with multidisciplinary approaches to disease etiology and health equity. We model trajectories as dynamical systems on the hyperbolic plane, representing epidemiological macrostates of population health burdens. Leveraging Global Burden of Disease (GBD) 2021 data for 10 major chronic conditions, we compute a statistic as the difference in total Fisher-Rao distances along male and female trajectories across the US, Western Europe, and Japan (1990–2019). We assess cross-regional consistency parametrically and nonparametrically, comparing against Kullback-Leibler divergence, absolute mean differences, and absolute standard deviation differences. This framework addresses novel issues in population-based epidemiology by providing a consistent, geometry-informed metric to characterise sex disparities in disease determinants and outcomes, enhancing public health surveillance and equity in chronic disease management.

## 2 Materials and Methods

### 2.1 Model Formulation

We developed a model motivated by hyperbolic dynamical systems theory [11, 12] to track changes in disease prevalence across five-year age groups stratified by sex. Dynamical systems provide a mathematical framework widely used in biology and medicine [13] to describe how state variables evolve over time or, as here, across successive age cohorts, analogous to cohort-progression models in epidemiology. To ensure prevalence rates remain realistic and non-negative, we apply a (base 10) logit transformation [14] and assume a normal distribution for the transformed data. Each epidemiological summary (in logit scale), or macrostate [15], represents the overall disease burden in a population subset, capturing its average level (mean, denoted *µ*) and variability (standard deviation, denoted *σ*). These macrostates are positioned on the hyperbolic plane, a curved geometric space suitable for statistical distributions because the metric has an information-theoretic interpretation [16, 17]:

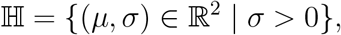

where ℍ denotes the hyperbolic plane in the Poincaré model, ℝ^2^ is two-dimensional real space, and *σ >* 0 ensures positive variability. In information geometry, the *statistical manifold* [18] of the normal distribution is defined as the set ℍ endowed with the *Fisher-Rao distance d*_FR_ [1, 2, 3], a metric that measures dissimilarity between gaussian random variables 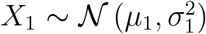 and 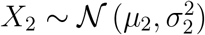, is given by the formula [19]:

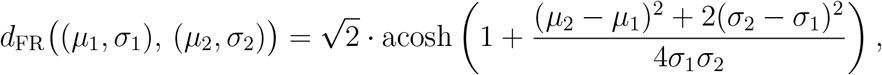

where acosh is the inverse hyperbolic cosine. The general definition of the Fisher-Rao distance [20], involving differential geometry, will not be used in the present article.

Age groups are defined in five-year intervals as 𝒜 = {*t*_*k*_ : *k* ∈ ℕ}, where *t*_*k*_ = [5*k*, 5*k* + 4] and ℕ denotes the natural numbers starting at 0. In practice, we consider *k* = 0, …, 18, corresponding to the age ranges 0–4 through 90–94 years. Biological sex categories are 𝒮 = {male, female}, consistent with the SAGER guidelines [21] and restricted to sex because the dataset does not include gender information [22]. The population space *X*, containing all possible populations, evolves under a mapping *f* : *X* → *X*, with iterations *x*_*n*+1_ = *f* (*x*_*n*_) representing aging 5 years, and the initial condition *x*_0_ = *x* ∈ *X*. The macrostate map *p* : *X* → ℍ assigns *p*(*x*) = (*µ, σ*). Subsets 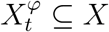 for sex *φ* and age *t* evolve as 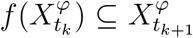. From Global Burden of Disease (GBD) 2021 data [22], we derive macrostate sequence *p*(*x*_0_), *p*(*x*_1_), …, *p*(*x*_18_) of *x*_0_, *x*_1_, …, *x*_18_, or disease prevalence trajectories, for each sex *φ*:

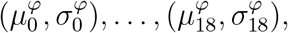

illustrated in Figures 1 and 2 for chronic respiratory diseases in California, 1990, by sex.

**Figure 1.**
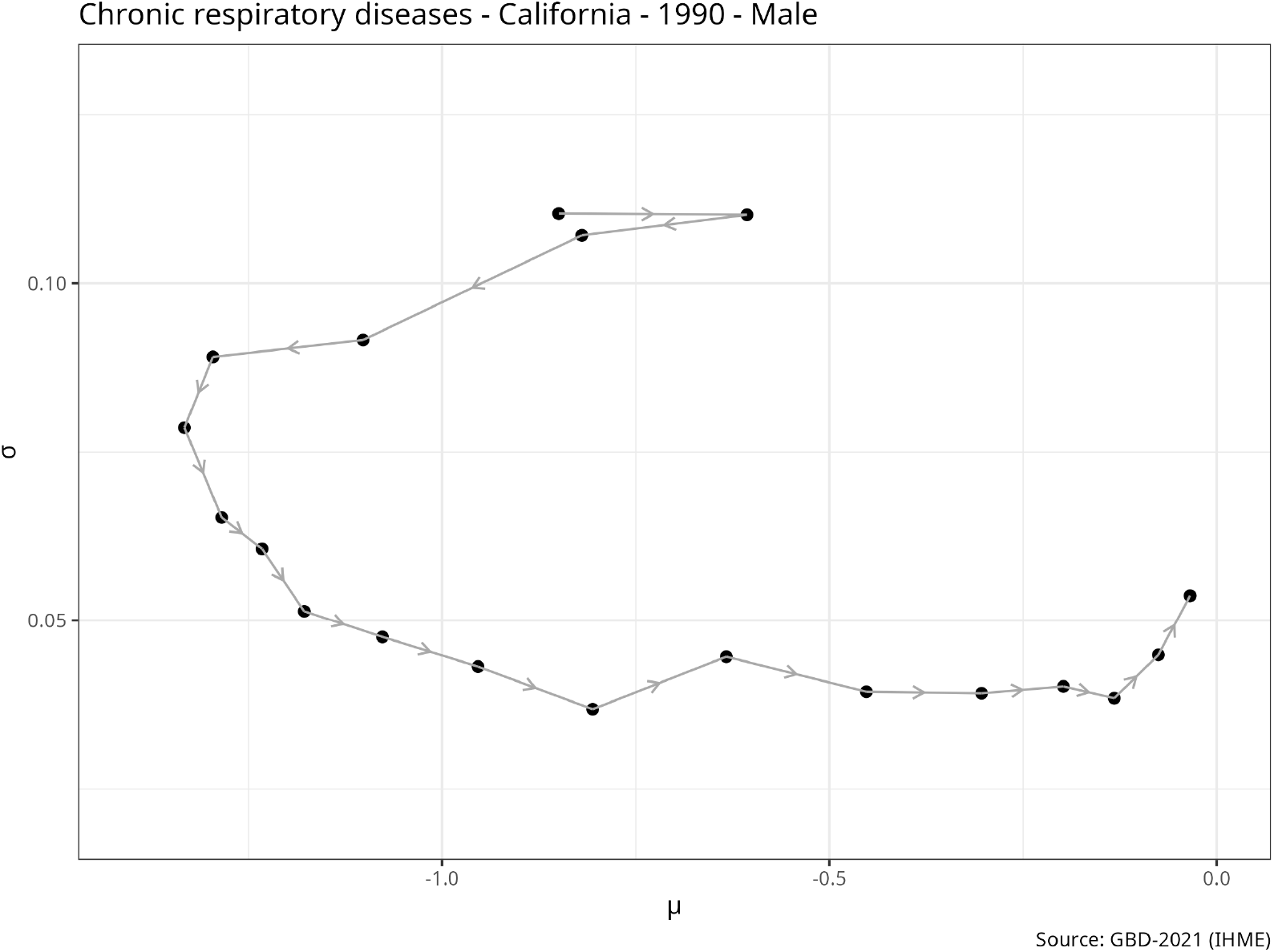
Trajectories in (*µ, σ*) space for chronic respiratory diseases in California (1990). Markers indicate successive time points (age groups); connecting lines show the temporal path of the parameter estimates. Sex: Male.

**Figure 2.**
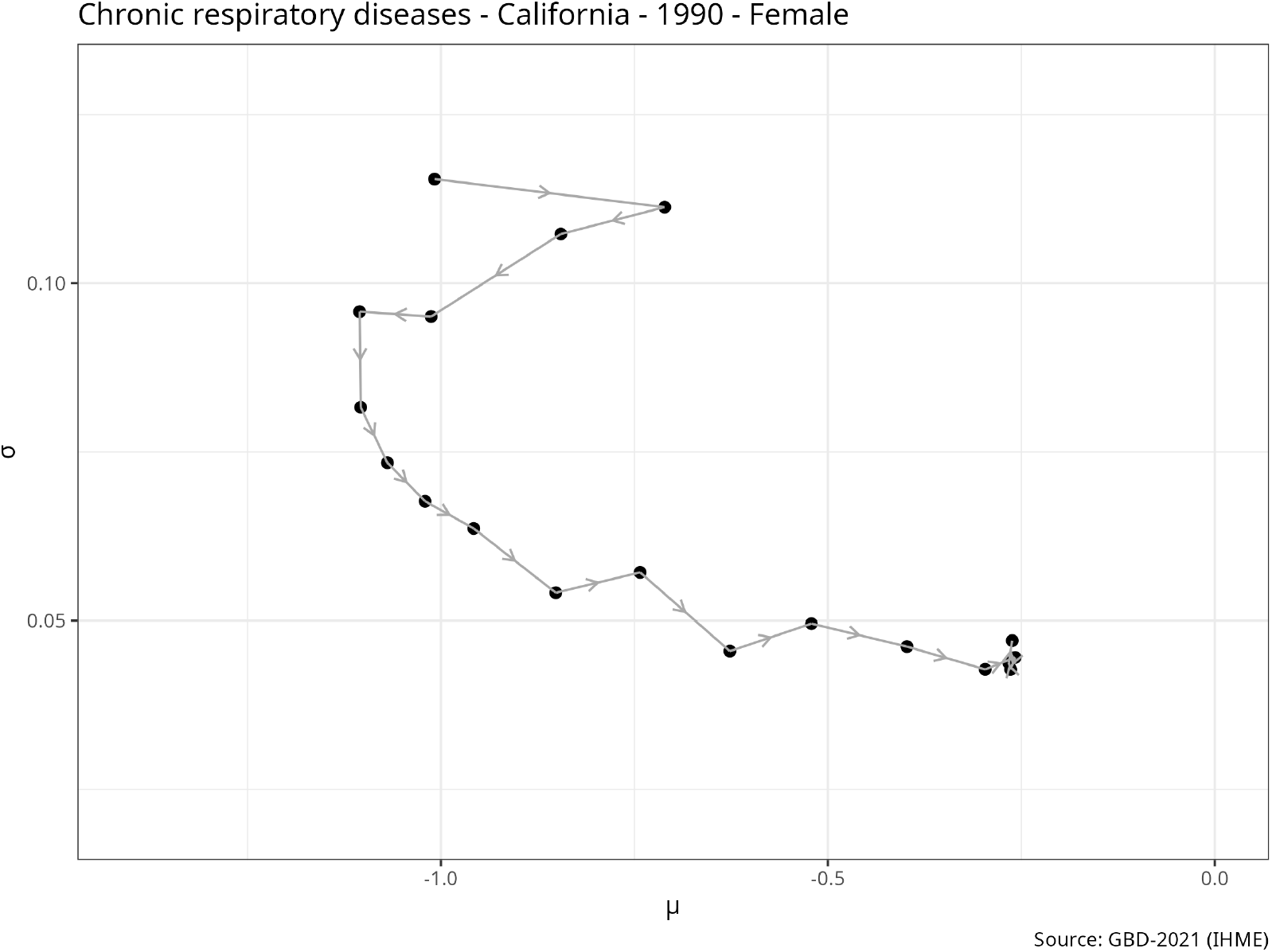
Trajectories in (*µ, σ*) space for chronic respiratory diseases in California (1990). Markers indicate successive time points (age groups); connecting lines show the temporal path of the parameter estimates. Sex: Female.

To reconcile the dynamical-systems framing with cross-sectional age slices we adopt a time-independent (autonomous) model for aging: the map *f* : *X* → *X* that advances a population state by one five-year step is assumed not to depend on calendar time. Equivalently, for any population state *x* and any calendar year *t* we assume the same aging rule applies, so that *x*_*n*+1_ = *f* (*x*_*n*_) has the same meaning whether it represents cohort *B* moved forward five years or cohort *A* observed five years older. Under this stationarity assumption a five-year older age slice in year *t* may be interpreted, within the model, as the same cohort observed in year *t* + 5, which permits treating cross-sectional age-profiles as synthetic cohort trajectories. We emphasize that this is a simplification: it ignores period effects (secular trends, epidemics, policy changes), cohort-specific exposures, selective mortality, migration, and other calendar-time heterogeneities that can make period age-profiles an imperfect proxy for true longitudinal cohort change. To assess robustness, we compare results across multiple calendar years.

### 2.2 Model Analysis

To quantify sex differences in prevalence evolution, we compute *d* = *d*^male^ − *d*^female^, where *d*^*φ*^ is the total trajectory length for sex *φ*:

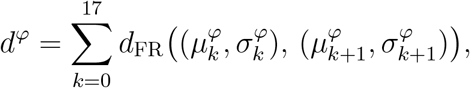

providing a geometrically meaningful notion of length for epidemiological shifts. We analyse *d* parametrically, assuming a normal distribution *d* ~ 𝒩 (*µ, σ*^2^) to estimate means and variances, and nonparametrically via the frequency of *d >* 0. Cumulative distances

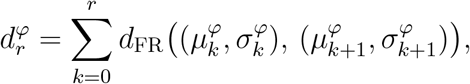

are plotted for intuition, as in Figure 3 for chronic respiratory diseases.

**Figure 3.**
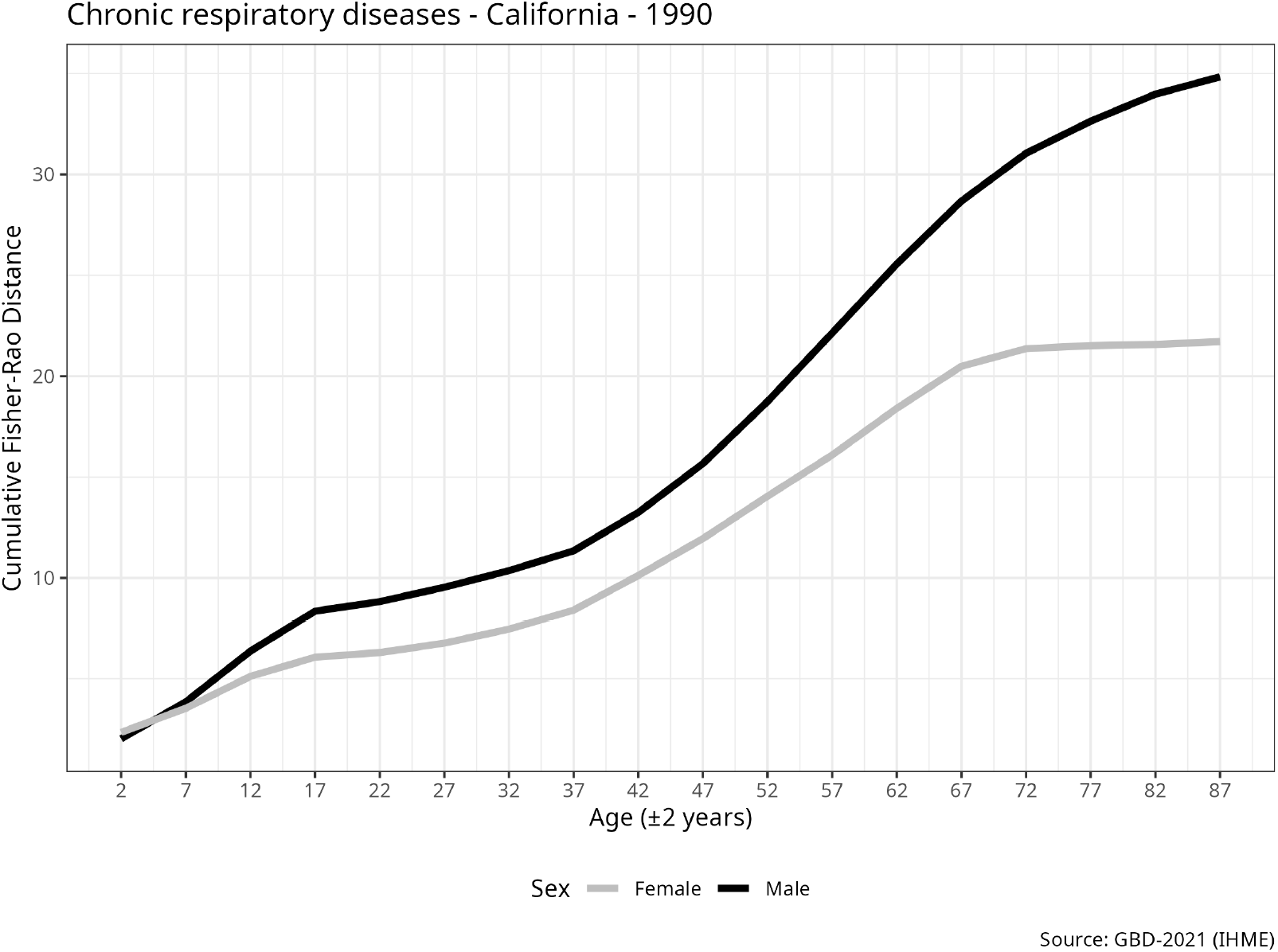
Cumulative Fisher-Rao distance 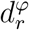 by sex for chronic respiratory diseases in California (1990). The horizontal axis is the centre 5*r* + 2 of the age group *t*_*r*_ =]5*r*, 5*r* + 4].

### 2.3 Fitting the model to data

We applied our model to data from the Global Burden of Disease Study, manually retrieved from the IHME website [22].

The R scripts used for data cleaning and analysis are provided in the supplementary material.

#### 2.3.1 Data Sources

Drawing on IHME’s GBD 2021 estimates [22, 23], we examine prevalence per 100,000, stratified by age, sex, location, and year, with 95% uncertainty intervals from systematic reviews and integrations [24, 25]. Focusing on the US (51 locations: 50 states and the District of Columbia), Western Europe (24 countries: Andorra, Austria, Belgium, Cyprus, Denmark, Finland, France, Germany, Greece, Iceland, Ireland, Israel, Italy, Luxembourg, Malta, Monaco, Netherlands, Norway, Portugal, San Marino, Spain, Sweden, Switzerland, United Kingdom), and Japan (47 prefectures), we compare age-specific trajectories to reveal sex differences, embracing the data’s open-ended nature over rigid unification.

#### 2.3.2 Data Extraction and Preprocessing

Age midpoints are assigned as 2 for *<*5, incrementing to 92 for 90–94. We logit-transform^1^ the prevalence assuming normality:

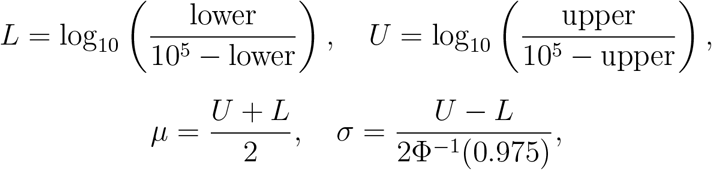

where Φ^−1^(0.975) ≈ 1.96 is the 97.5th percentile point of the standard normal distribution.

Processed CSV files—one per region—contain age, sex, disease, subregional location, year, and the parameters *µ* and *σ*. Data from the pandemic years 2020–2021 were excluded to mitigate bias [26].

### 2.4 Cross-Regional Consistency

We evaluated the consistency of sex-specific shifts in age-stratified disease prevalence trajectories across the United States, Western Europe, and Japan from 1990 to 2019, using four metrics to quantify total trajectory shifts: absolute mean difference (*µ*-distance)

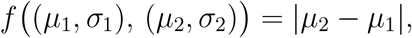

absolute standard deviation difference (*σ*-distance)

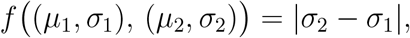

Kullback-Leibler divergence (KL-divergence) [27]

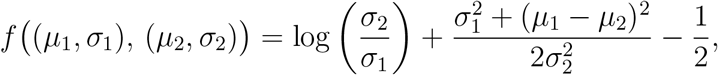

and Fisher-Rao distance (FR-distance)

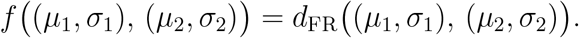

For each sex *φ*, disease, year, and location, the total shift

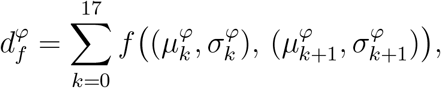

was the sum of distances across 18 consecutive 5-year age groups, assuming normal distributions for prevalence parameters.

In the parametric approach, sex differences were computed as 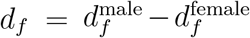 per location (inside a region), then averaged to estimate mean *µ* per region, assuming normality of *d*_*f*_. Consistency was defined as the number of diseases (out of 10) with uniform sign of *µ* across all regions (all positive or all negative). If *µ* = 0 in any region, it is interpreted as an inconsistency, as zero has no sign.

In the nonparametric approach, Cohen’s *g* measured the probability of greater male shifts minus 0.5 across locations per region, i.e.,

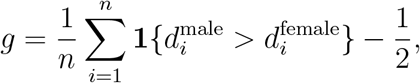

where *n* is the number of locations and **1**{·} is the indicator function. According to Jacob Cohen [28], the effect size *g* is conventionally interpreted as small, medium, or large when |*g* ≈ |0.05, |*g*| ≈ 0.15, and |*g*|≈ 0.25, respectively.

Consistency was the number of diseases with uniform sign of *g* across regions (all positive or all negative). If *g* = 0 in any region, it is interpreted as an inconsistency, as zero has no sign.

### 2.5 Sex-Specific Shifts

Using FR-distance, we computed sex differences *d* = *d*^male^ − *d*^female^ per location, then derived regional summary statistics: Cohen’s *g*, mean *µ*, standard deviation *σ*, skewness *γ*_1_, excess kurtosis *κ*, Shapiro-Wilk p-value *p*_*SW*_ for normality, and t-test p-value *p*_*t*_ for mean deviation from zero. Results for 1990 exemplify patterns across 1990–2019.

It is important to note the ambiguity in the notation: the prevalence is modelled as being normally distributed on the base-10 logit scale, 𝒩 (*µ, σ*^2^), whereas in the parametric model, we assume *d* ~ 𝒩 (*µ, σ*^2^). However, the parameters *µ* and *σ* in these two contexts are not the same.

### 2.6 Ethics Statement

This study uses publicly available secondary data from IHME GBD 2021 [22], exempt from ethical review per institutional guidelines and the Declaration of Helsinki. No human subjects were involved.

## 3 Results

### 3.1 Cross-Regional Consistency

Supplementary table for parametric consistency (1990–2019) show FR-distance yielded the highest consistency (e.g., 10/10 diseases in early 1990s).

Supplementary table for nonparametric consistency (1990–2019) confirms FR-distance’s superior performance (e.g., 10/10 in mid-1990s).

These findings highlight FR-distance’s utility in capturing geodesic shifts on the hyperbolic plane of normal distribution parameters, offering a novel epidemiologic method for assessing trajectory consistency in population health data.

### 3.2 Sex-Specific Shifts

Diseases with consistently greater male shifts (positive *g* and *µ, p*_*t*_ *<* 0.05) included neoplasms (supplementary table showing *g* = 0.50, *µ* = 11.44 to 17.61 across regions), cardiovascular diseases (supplementary table showing *g* = 0.27 to 0.50, *µ* = 0.69 to 4.87), chronic respiratory diseases (supplementary table showing *g* = 0.21 to 0.50, *µ* = 1.77 to 13.21), diabetes and kidney diseases (supplementary table showing *g* = 0.42 to 0.50, *µ* = 1.82 to 3.91), skin and subcutaneous diseases (supplementary table showing *g* = 0.50, *µ* = 5.61 to 12.14), and sense organ diseases (supplementary table showing *g* = 0.50, *µ* = 3.79 to 6.92).

Diseases with greater female shifts (negative *g* and *µ, p*_*t*_ *<* 0.05) included neurological disorders (supplementary table showing *g* = −0.50, *µ* = −0.86 to −2.78), mental disorders (supplementary table showing *g* = −0.21 to −0.50, *µ* = −0.42 to −6.48), and substance use disorders (supplementary table showing *g* = −0.42 to −0.50, *µ* = −1.56 to −3.28).

Digestive diseases showed mixed patterns, with no significant sex differences in the United States (*p*_*t*_ = 0.19) or Western Europe (*p*_*t*_ = 0.63), but male predominance in Japan (*p*_*t*_ *<* 0.05; supplementary table showing *g* = 0.00 to 0.31, *µ* = 0.07 to 0.54).

This application of FR-distance provides a robust, geometry-informed metric for quantifying sex disparities in disease trajectories, enhancing etiologic insights and equity assessments in public health surveillance.

## 4 Discussion

This study introduces a novel application of the Fisher-Rao geodesic distance, rooted in information geometry, to quantify sex-based differences in disease prevalence trajectories across age groups using GBD 2021 data from 1990 to 2019. By modelling trajectories as dynamical systems on the hyperbolic plane, the metric provides superior cross-regional consistency compared to alternatives like Kullback-Leibler divergence, absolute mean differences, and absolute standard deviation differences. Key findings reveal greater male shifts in neoplasms, cardiovascular diseases, chronic respiratory diseases, diabetes and kidney diseases, skin and subcutaneous diseases, and sense organ diseases; greater female shifts in neurological disorders, mental disorders, and substance use disorders; and variable patterns for digestive diseases across the US, Western Europe, and Japan.

These results advance epidemiologic methods by offering a geometry-informed tool to assess sex disparities in chronic disease burdens, enhancing public health surveillance and equity assessments. The framework’s consistency supports its use in population-based analyses of disease determinants, aligning with multidisciplinary approaches to etiology.

Limitations include data biases from the COVID-19 pandemic—elevated prevalence for major depressive and anxiety disorders, unchanged self-harm rates [26]—prompting exclusion of 2020–2021 data, and lack of gender information, limiting focus to biological sex.

Gender, a multifaceted social determinant, influences health via environmental exposures like air pollution [29] and requires integration of gender-affirming hormone therapy in frameworks [30]. To address its complexity, avoid binary models; instead, represent gender as a vector space capturing cultural, social, and relational dimensions using Likert scales [31, 32, 33, 34]. Future extensions include applying Fisher-Rao distance to gender-based differences, including transgender population, formalising population dynamics to prove theorems for particular models (without aiming to unify biosciences [35]), and developing a notion of entropy [36] to answer the question how many microstates (possible populations) correspond to the same macrostate (*µ, σ*)?

## 5 Conclusions

This paper introduces an innovative Fisher-Rao geodesic distance framework that models disease prevalence as trajectories on the hyperbolic plane to measure sex-based disparities between age groups with greater cross-regional consistency. Applied to GBD 2021 data for 10 major chronic disease categories in the US, Western Europe, and Japan (1990–2019), it outperforms Kullback-Leibler divergence and absolute mean/standard deviation differences. Males show greater shifts in neoplasms, cardiovascular, chronic respiratory, diabetes/kidney, skin/subcutaneous, and sense organ diseases; females in neurological, mental, and substance use disorders; digestive diseases vary by region. This geometry-informed metric advances epidemiologic methods for assessing disease burden determinants, promoting health equity through population-based analyses of sex differences. Future work should integrate gender dimensions, including transgender populations, to enhance public health surveillance and address inequities.

## Supporting information

Code in R to reproduce the results

## Data Availability

All data produced are available online at
Global Burden of Disease Study 2021 (GBD 2021)
https://ghdx.healthdata.org/gbd-2021

## Abbreviations

GBD: Global Burden of Disease
IHME: Institute for Health Metrics and Evaluation
FR: Fisher-Rao
KL: Kullback-Leibler
SAGER: Sex and Gender Equity in Research

## 6 Acknowledgements

None. Funding: None declared. Competing Interests: None declared.

## 7 Declaration of generative AI and AI-assisted technologies in the writing process

During the preparation of this work the author used Grok 4 Heavy for language editing, LaTeX, R code, bibliographic suggestions and brainstorming. The author reviewed and edited the content as needed and takes full responsibility for the content of the published article.

## 8 Data availability

The data is available on the website: https://vizhub.healthdata.org/gbd-results/

## A Tables in Parametric Approach

**Table 1:**
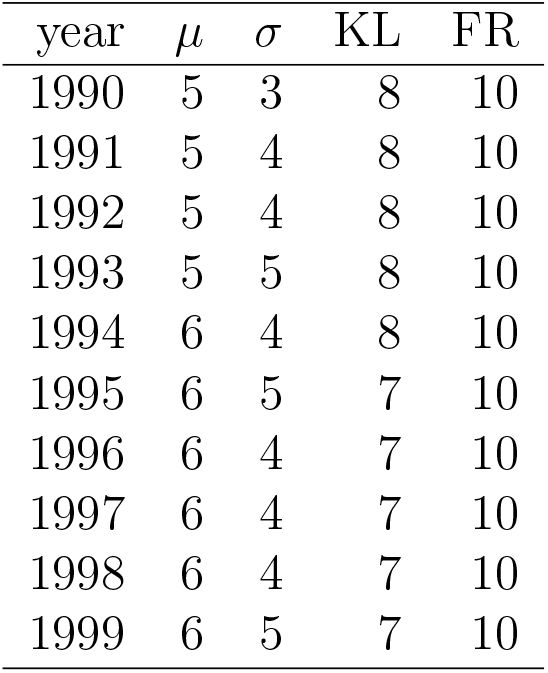
Consistency:1990–1999 (parametric)

**Table 2:**
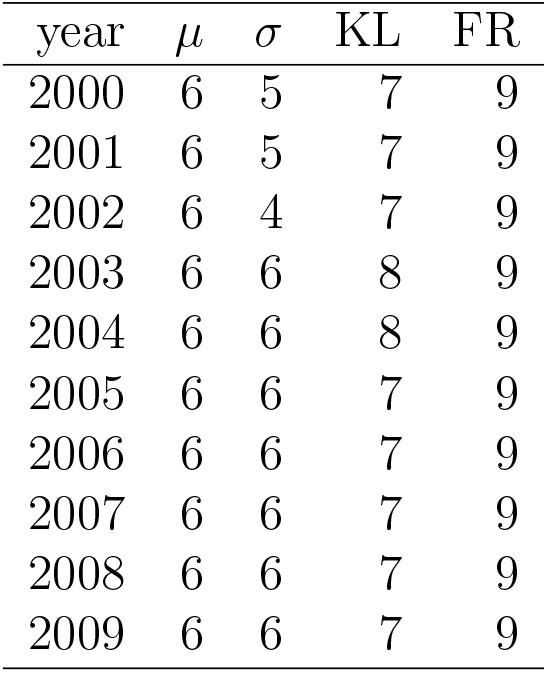
Consistency: 2000–2009 (parametric)

**Table 3:**
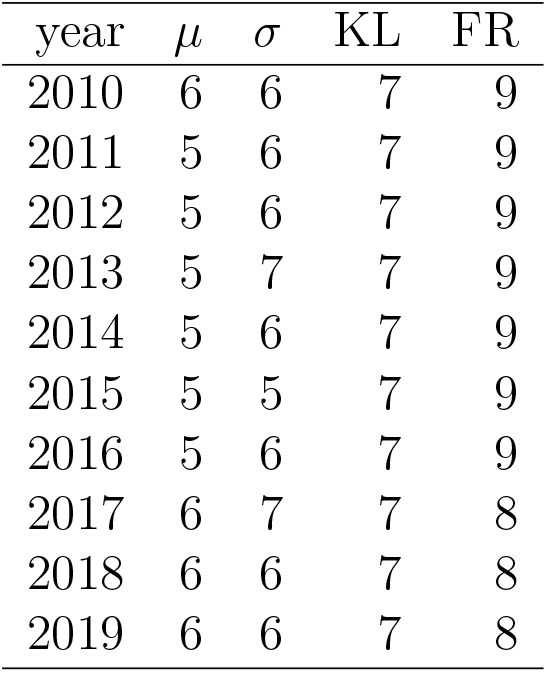
Consistency: 2010–2019 (parametric)

## B Tables in Nonparametric Approach

**Table 4:**
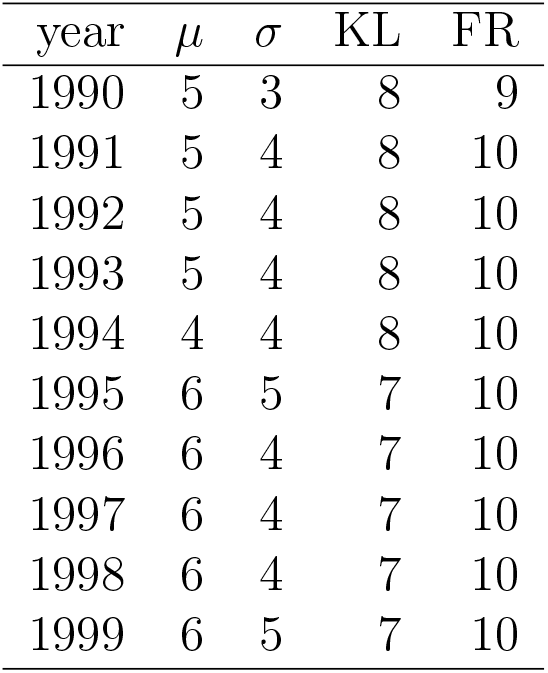
Consistency: 1990–1999 (nonparametric)

**Table 5:**
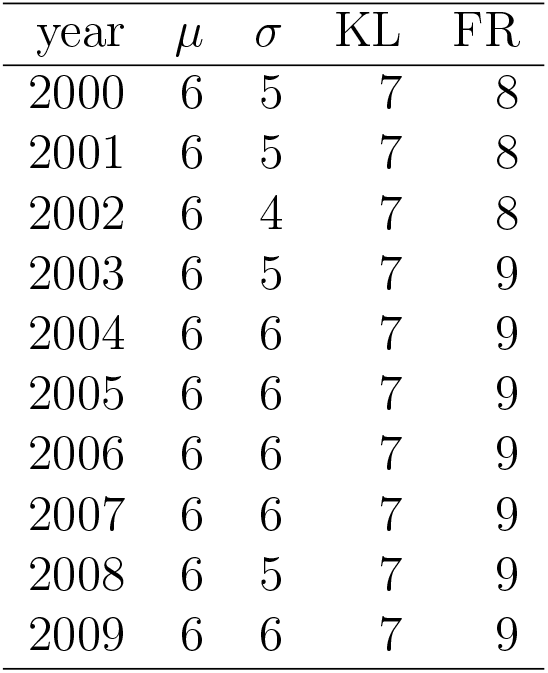
Consistency: 2000–2009 (nonparametric)

**Table 6:**
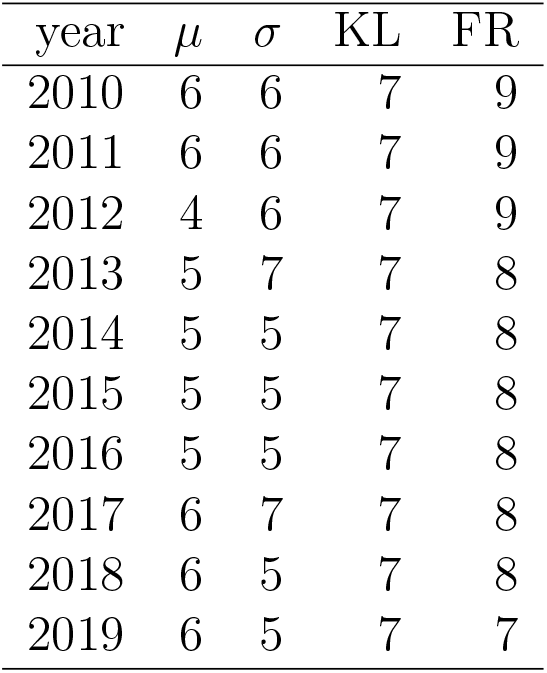
Consistency: 2010–2019 (nonparametric)

## C Tables in Diseases with Greater Male Shifts

**Table 7:** Neoplasms parameters (greater male shifts). Year: 1990.

| region | $g$ | $\mu$ | $\sigma$ | $\gamma_1$ | $\kappa$ | $p_{SW}$ | $p_t$ |
| --- | --- | --- | --- | --- | --- | --- | --- |
| United States | 0.50 | 11.44 | 1.38 | 0.90 | 3.59 | 0.00 | 0.00 |
| Western Europe | 0.50 | 17.61 | 5.12 | -0.38 | -0.43 | 0.68 | 0.00 |
| Japan | 0.50 | 15.97 | 1.49 | -0.00 | -0.03 | 0.88 | 0.00 |

**Table 8:** Cardiovascular diseases parameters (greater male shifts). Year: 1990.

| region | $g$ | $\mu$ | $\sigma$ | $\gamma_1$ | $\kappa$ | $p_{SW}$ | $p_t$ |
| --- | --- | --- | --- | --- | --- | --- | --- |
| United States | 0.42 | 1.23 | 0.78 | -0.30 | -0.40 | 0.46 | 0.00 |
| Western Europe | 0.50 | 4.87 | 2.39 | -0.25 | -0.84 | 0.75 | 0.00 |
| Japan | 0.27 | 0.69 | 1.12 | -0.47 | 0.01 | 0.24 | 0.00 |

**Table 9:**
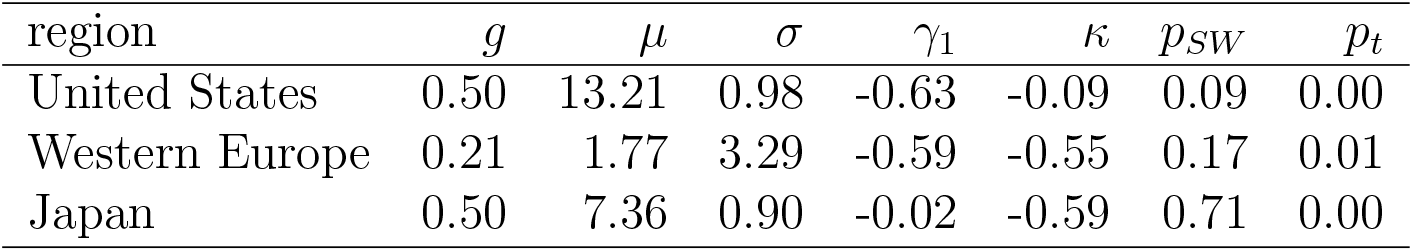
Chronic respiratory diseases parameters (greater male shifts). Year: 1990.

| region | $g$ | $\mu$ | $\sigma$ | $\gamma_1$ | $\kappa$ | $p_{SW}$ | $p_t$ |
| --- | --- | --- | --- | --- | --- | --- | --- |
| United States | 0.50 | 13.21 | 0.98 | -0.63 | -0.09 | 0.09 | 0.00 |
| Western Europe | 0.21 | 1.77 | 3.29 | -0.59 | -0.55 | 0.17 | 0.01 |
| Japan | 0.50 | 7.36 | 0.90 | -0.02 | -0.59 | 0.71 | 0.00 |

**Table 10:**
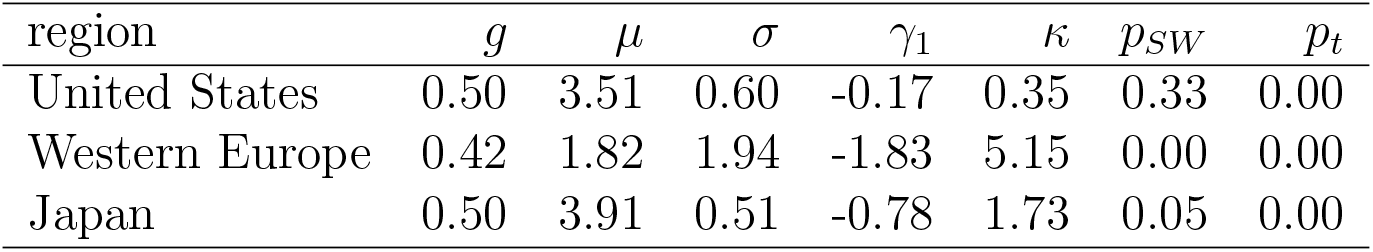
Diabetes and kidney diseases parameters (greater male shifts). Year: 1990.

**Table 11:** Skin and subcutaneous diseases parameters (greater male shifts). Year: 1990.

| region | $g$ | $\mu$ | $\sigma$ | $\gamma_1$ | $\kappa$ | $p_{SW}$ | $p_t$ |
| --- | --- | --- | --- | --- | --- | --- | --- |
| United States | 0.50 | 12.14 | 1.97 | 0.06 | -0.74 | 0.81 | 0.00 |
| Western Europe | 0.50 | 5.61 | 0.80 | 0.81 | 0.06 | 0.05 | 0.00 |
| Japan | 0.50 | 6.93 | 0.38 | 0.19 | -0.50 | 0.86 | 0.00 |

**Table 12:**
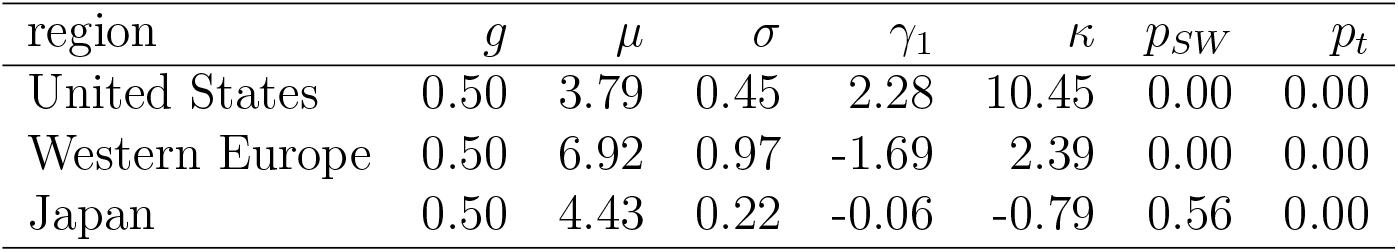
Sense organ diseases parameters (greater male shifts). Year: 1990.

| region | $g$ | $\mu$ | $\sigma$ | $\gamma_1$ | $\kappa$ | $p_{SW}$ | $p_t$ |
| --- | --- | --- | --- | --- | --- | --- | --- |
| United States | 0.50 | 3.79 | 0.45 | 2.28 | 10.45 | 0.00 | 0.00 |
| Western Europe | 0.50 | 6.92 | 0.97 | -1.69 | 2.39 | 0.00 | 0.00 |
| Japan | 0.50 | 4.43 | 0.22 | -0.06 | -0.79 | 0.56 | 0.00 |

## D Tables in Diseases with Greater Female Shifts

**Table 13:** Neurological disorders parameters (greater female shifts). Year: 1990.

| region | $g$ | $\mu$ | $\sigma$ | $\gamma_1$ | $\kappa$ | $p_{SW}$ | $p_t$ |
| --- | --- | --- | --- | --- | --- | --- | --- |
| United States | -0.50 | -1.45 | 0.17 | 0.05 | 1.83 | 0.07 | 0.00 |
| Western Europe | -0.50 | -2.78 | 0.44 | -1.93 | 5.14 | 0.00 | 0.00 |
| Japan | -0.50 | -0.86 | 0.19 | 0.09 | 0.43 | 0.50 | 0.00 |

**Table 14:** Mental disorders parameters (greater female shifts). Year: 1990.

| region | $g$ | $\mu$ | $\sigma$ | $\gamma_1$ | $\kappa$ | $p_{SW}$ | $p_t$ |
| --- | --- | --- | --- | --- | --- | --- | --- |
| United States | -0.50 | -6.48 | 0.43 | -0.29 | 0.73 | 0.16 | 0.00 |
| Western Europe | -0.21 | -0.42 | 0.90 | -0.84 | 0.38 | 0.02 | 0.03 |
| Japan | -0.50 | -1.00 | 0.41 | -0.83 | 4.02 | 0.00 | 0.00 |

**Table 15:** Substance use disorders parameters (greater female shifts). Year: 1990.

| region | $g$ | $\mu$ | $\sigma$ | $\gamma_1$ | $\kappa$ | $p_{SW}$ | $p_t$ |
| --- | --- | --- | --- | --- | --- | --- | --- |
| United States | -0.50 | -3.28 | 1.12 | -0.29 | -0.56 | 0.38 | 0.00 |
| Western Europe | -0.42 | -2.82 | 1.95 | -0.06 | -0.56 | 0.96 | 0.00 |
| Japan | -0.50 | -1.56 | 0.41 | 0.34 | -0.45 | 0.37 | 0.00 |

## E Table in Mixed Patterns

**Table 16:**
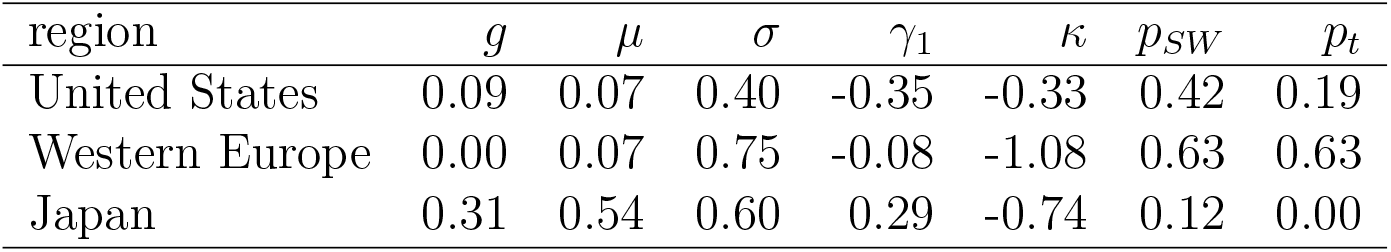
Digestive diseases parameters (mixed patterns). Year: 1990. Note that the fact *g* = 0 in the Western European data corresponds to an inconsistency of the Fisher-Rao distance according to our definition of consistency.

## Footnotes

1 We use base-10 logarithms for intuition, reflecting the ubiquity of the decimal system.

