## Supplementary material for "Fisher-Rao distance and sex differences in disease prevalence trajectories": Code in R to reproduce the results: code_1.R

```
logit_per_100k_aux <- function(x) {
  log10(x) - log10(1e5 - x)
}

logit_per_100k <- function(x) {
  sapply(x, function(y) logit_per_100k_aux(y))
}

is_numeric_age <- function(age) {
  accepted_formats <- c(
    "<5 years",
    "5-9 years",
    "10-14 years",
    "15-19 years",
    "20-24 years",
    "25-29 years",
    "30-34 years",
    "35-39 years",
    "40-44 years",
    "45-49 years",
    "50-54 years",
    "55-59 years",
    "60-64 years",
    "65-69 years",
    "70-74 years",
    "75-79 years",
    "80-84 years",
    "85-89 years",
    "90-94 years"
  )
  age %in% accepted_formats
}

number_from_age <- function(age) {
  c(
    "<5 years" = 2,
    "5-9 years" = 7,
    "10-14 years" = 12,
    "15-19 years" = 17,
    "20-24 years" = 22,
    "25-29 years" = 27,
    "30-34 years" = 32,
    "35-39 years" = 37,
    "40-44 years" = 42,
    "45-49 years" = 47,
    "50-54 years" = 52,
    "55-59 years" = 57,
    "60-64 years" = 62,
    "65-69 years" = 67,
    "70-74 years" = 72,
    "75-79 years" = 77,
    "80-84 years" = 82,
    "85-89 years" = 87,
    "90-94 years" = 92
  )[age]
}

get_mu_sigma_aux <- function(df, k=0.255106728462327014828) {
  df |> dplyr::mutate(
    mu = 0.5*(Upper + Lower),
    sigma = k*(Upper - Lower)
  ) |>
    dplyr::select(-Lower, -Upper)
}

get_mu_sigma <- function(df) {
  df |> 
    dplyr::mutate(
      Lower = logit_per_100k(lower),
      Upper = logit_per_100k(upper)
  ) |>
    dplyr::select(-lower, -upper) |>
    get_mu_sigma_aux()
}

data_cleaning <- function(df) {
  df |>
    dplyr::filter(year >= 1990, year <= 2019) |>
    dplyr::select(-val) |>
    dplyr::filter(is_numeric_age(age)) |>
    dplyr::mutate(
      disease = cause,
      age = as.integer(number_from_age(age)),
      year = as.integer(year)
  ) |>
    get_mu_sigma() |>
    dplyr::select(age, sex, disease, location, year, mu, sigma)
}

read_file <- function(filename, measure, disease, country) {
  readr::read_csv(fs::path(here::here("data", "IHME-GBD_2021", 
              measure, country, disease, filename, filename),
                           ext = "csv"))
}

read_country_clean <- function(files, measure, disease, country) {
  purrr::map_dfr(
    .x = files,
    .f = function(file) 
      read_file(filename = file,
                measure = measure,
                disease = disease,
                country = country) |> 
      data_cleaning()
  )
}

main <- function(disease_list, output_name, country) {
  purrr::map_dfr(
    .x = names(disease_list),
    .f = function(disease_name) 
      read_country_clean( 
        files=disease_list[[disease_name]],
        measure = "prevalence",
        disease=disease_name ,
        country = country
      )
  ) |>
    readr::write_csv(
      file = fs::path(here::here("output", "code_1", 
                                 paste0(output_name,"-",country)), ext="csv")
      )
}


main(
  disease_list = list(
    "Neoplasms" = c("IHME-GBD_2021_DATA-0ccfabe8-1",
                    "IHME-GBD_2021_DATA-4de97570-1",
                    "IHME-GBD_2021_DATA-5e9c5f7f-1",
                    "IHME-GBD_2021_DATA-4541a005-1",
                    "IHME-GBD_2021_DATA-bf5e053f-1",
                    "IHME-GBD_2021_DATA-ce23996a-1",
                    "IHME-GBD_2021_DATA-f4ad4758-1"),
    "Cardiovascular diseases" = c("IHME-GBD_2021_DATA-2c7e75f7-1",
                                  "IHME-GBD_2021_DATA-9f2f5a44-1",
                                  "IHME-GBD_2021_DATA-91a3e118-1",
                                  "IHME-GBD_2021_DATA-201ef8ca-1",
                                  "IHME-GBD_2021_DATA-2809e7aa-1",
                                  "IHME-GBD_2021_DATA-c1f79489-1",
                                  "IHME-GBD_2021_DATA-ee37dc9c-1"),
    "Chronic respiratory diseases" = c("IHME-GBD_2021_DATA-2c0b2e99-1",
                                       "IHME-GBD_2021_DATA-6d489555-1",
                                       "IHME-GBD_2021_DATA-181e8b0f-1",
                                       "IHME-GBD_2021_DATA-a5918879-1",
                                       "IHME-GBD_2021_DATA-cd3e6aa4-1",
                                       "IHME-GBD_2021_DATA-e978da51-1",
                                       "IHME-GBD_2021_DATA-fd586fb9-1"),
    "Digestive diseases" = c("IHME-GBD_2021_DATA-0c662824-1",
                             "IHME-GBD_2021_DATA-8c8ce4b5-1",
                             "IHME-GBD_2021_DATA-9fe35a8e-1",
                             "IHME-GBD_2021_DATA-b78dc952-1",
                             "IHME-GBD_2021_DATA-c1a79723-1",
                             "IHME-GBD_2021_DATA-d30a7bb8-1",
                             "IHME-GBD_2021_DATA-e32a1f4c-1"),
    "Neurological disorders" = c("IHME-GBD_2021_DATA-2b014eb8-1",
                                 "IHME-GBD_2021_DATA-67258e73-1",
                                 "IHME-GBD_2021_DATA-99424683-1",
                                 "IHME-GBD_2021_DATA-ae5e7398-1",
                                 "IHME-GBD_2021_DATA-c528a786-1",
                                 "IHME-GBD_2021_DATA-e8bd8ef4-1",
                                 "IHME-GBD_2021_DATA-f164718c-1"),
    "Mental disorders" = c("IHME-GBD_2021_DATA-9cc85bc6-1",
                           "IHME-GBD_2021_DATA-58e10194-1",
                           "IHME-GBD_2021_DATA-88f1c4a9-1",
                           "IHME-GBD_2021_DATA-a68d95ab-1",
                           "IHME-GBD_2021_DATA-d8a9ec46-1",
                           "IHME-GBD_2021_DATA-d20ef9a3-1",
                           "IHME-GBD_2021_DATA-e297ba96-1"),
    "Substance use disorders" = c("IHME-GBD_2021_DATA-0d353030-1",
                                  "IHME-GBD_2021_DATA-8e19d386-1",
                                  "IHME-GBD_2021_DATA-55ec3a78-1",
                                  "IHME-GBD_2021_DATA-633f5d90-1",
                                  "IHME-GBD_2021_DATA-725763ca-1",
                                  "IHME-GBD_2021_DATA-eb1fde78-1",
                                  "IHME-GBD_2021_DATA-fe58e123-1"),
    "Diabetes and kidney diseases" = c("IHME-GBD_2021_DATA-a251d1d6-1",
                                       "IHME-GBD_2021_DATA-9c09b3e0-1",
                                       "IHME-GBD_2021_DATA-76ae28cd-1",
                                       "IHME-GBD_2021_DATA-927af190-1",
                                       "IHME-GBD_2021_DATA-4997bf27-1",
                                       "IHME-GBD_2021_DATA-dc0632ae-1",
                                       "IHME-GBD_2021_DATA-edcea618-1"),
    "Skin and subcutaneous diseases" = c("IHME-GBD_2021_DATA-588af0ee-1",
                                         "IHME-GBD_2021_DATA-2731bf21-1",
                                         "IHME-GBD_2021_DATA-9807dd29-1",
                                         "IHME-GBD_2021_DATA-51110b36-1",
                                         "IHME-GBD_2021_DATA-45336123-1",
                                         "IHME-GBD_2021_DATA-ba8dae7a-1",
                                         "IHME-GBD_2021_DATA-cd010248-1"),
    "Sense organ diseases" = c("IHME-GBD_2021_DATA-0ec8404f-1",
                               "IHME-GBD_2021_DATA-5f76094f-1",
                               "IHME-GBD_2021_DATA-6d030537-1",
                               "IHME-GBD_2021_DATA-44a41a41-1",
                               "IHME-GBD_2021_DATA-62d4a2be-1",
                               "IHME-GBD_2021_DATA-77967b25-1",
                               "IHME-GBD_2021_DATA-d1c9c886-1")
  ),
  output_name = "IHME-GBD_2021_CLEAN_prevalence",
  country = "Japan"
)


main(
  disease_list = list(
    "Neoplasms" = c("IHME-GBD_2021_DATA-6ecc1488-1",
                    "IHME-GBD_2021_DATA-1378a3ff-1",
                    "IHME-GBD_2021_DATA-bb448ff7-1",
                    "IHME-GBD_2021_DATA-ed8d2db3-1"),
    "Cardiovascular diseases" = c("IHME-GBD_2021_DATA-079a8e44-1",
                                  "IHME-GBD_2021_DATA-183fd27b-1",
                                  "IHME-GBD_2021_DATA-867319a1-1",
                                  "IHME-GBD_2021_DATA-d6de8a03-1",
                                  "IHME-GBD_2021_DATA-5320f34a-1"),
    "Chronic respiratory diseases" = c("IHME-GBD_2021_DATA-24aecd43-1",
                                       "IHME-GBD_2021_DATA-254f7ff3-1",
                                       "IHME-GBD_2021_DATA-440a3c63-1",
                                       "IHME-GBD_2021_DATA-a7e45a71-1"),
    "Digestive diseases" = c("IHME-GBD_2021_DATA-2a1a4e2b-1",
                             "IHME-GBD_2021_DATA-42ec3b32-1",
                             "IHME-GBD_2021_DATA-885c2056-1",
                             "IHME-GBD_2021_DATA-b3e65a6b-1",
                             "IHME-GBD_2021_DATA-5fdfa379-1"),
    "Neurological disorders" = c("IHME-GBD_2021_DATA-4ad1d6ca-1",
                                 "IHME-GBD_2021_DATA-9fb87864-1",
                                 "IHME-GBD_2021_DATA-22e1390f-1",
                                 "IHME-GBD_2021_DATA-ee54cf47-1"),
    "Mental disorders" = c("IHME-GBD_2021_DATA-1c88dabb-1",
                           "IHME-GBD_2021_DATA-87f5c2d1-1",
                           "IHME-GBD_2021_DATA-bbb7ae19-1",
                           "IHME-GBD_2021_DATA-f289d6f3-1",
                           "IHME-GBD_2021_DATA-42c22bca-1"),
    "Substance use disorders" = c("IHME-GBD_2021_DATA-4b0f8d27-1",
                                  "IHME-GBD_2021_DATA-a5cd1043-1",
                                  "IHME-GBD_2021_DATA-c9850cf1-1",
                                  "IHME-GBD_2021_DATA-fcdb4249-1"),
    "Diabetes and kidney diseases" = c("IHME-GBD_2021_DATA-7a165380-1",
                                       "IHME-GBD_2021_DATA-be7bfd85-1",
                                       "IHME-GBD_2021_DATA-f1c53142-1",
                                       "IHME-GBD_2021_DATA-fff67d3c-1"),
    "Skin and subcutaneous diseases" = c("IHME-GBD_2021_DATA-6e349f5b-1",
                                         "IHME-GBD_2021_DATA-565f2034-1",
                                         "IHME-GBD_2021_DATA-cc2b6d7b-1",
                                         "IHME-GBD_2021_DATA-f224b084-1"),
    "Sense organ diseases" = c("IHME-GBD_2021_DATA-6cea1c53-1",
                               "IHME-GBD_2021_DATA-086efe43-1",
                               "IHME-GBD_2021_DATA-402dcd0c-1",
                               "IHME-GBD_2021_DATA-a992c7f1-1",
                               "IHME-GBD_2021_DATA-2461708f-1")
    
    ),
  output_name = "IHME-GBD_2021_CLEAN_prevalence",
  country = "Western Europe"
)


main(
  disease_list = list(
    "Neoplasms" = c("IHME-GBD_2021_DATA-2c1986ea-1",
                    "IHME-GBD_2021_DATA-235ee5f8-1", 
                    "IHME-GBD_2021_DATA-4224b4e6-1",
                    "IHME-GBD_2021_DATA-5830439f-1",
                    "IHME-GBD_2021_DATA-ae4634a8-1",
                    "IHME-GBD_2021_DATA-b8a89a15-1",
                    "IHME-GBD_2021_DATA-d454da6b-1"),
    "Cardiovascular diseases" = c("IHME-GBD_2021_DATA-8bdda43a-1",
                                  "IHME-GBD_2021_DATA-47a6a283-1",
                                  "IHME-GBD_2021_DATA-72dec9d2-1",
                                  "IHME-GBD_2021_DATA-594c2fa6-1",
                                  "IHME-GBD_2021_DATA-76207db5-1",
                                  "IHME-GBD_2021_DATA-b5a965be-1",
                                  "IHME-GBD_2021_DATA-fee1d443-1"
    ),
    "Chronic respiratory diseases" = c("IHME-GBD_2021_DATA-8d7180df-1",
             "IHME-GBD_2021_DATA-032c907b-1",
             "IHME-GBD_2021_DATA-77da5abb-1",
             "IHME-GBD_2021_DATA-86bd88fa-1", 
             "IHME-GBD_2021_DATA-43538444-1",
             "IHME-GBD_2021_DATA-a2e0c894-1",
             "IHME-GBD_2021_DATA-ef3a720a-1"),
    "Digestive diseases" = c("IHME-GBD_2021_DATA-0cd6e564-1",
                             "IHME-GBD_2021_DATA-6ae43e37-1",
                             "IHME-GBD_2021_DATA-51c86264-1",
                             "IHME-GBD_2021_DATA-a1785b2e-1",
                             "IHME-GBD_2021_DATA-ae1b269a-1",
                             "IHME-GBD_2021_DATA-d2ac8965-1",
                             "IHME-GBD_2021_DATA-e0221152-1",
                             "IHME-GBD_2021_DATA-72c9395a-1"),
    "Neurological disorders" = c("IHME-GBD_2021_DATA-30eccdb0-1", 
                                 "IHME-GBD_2021_DATA-35cca385-1",
                                 "IHME-GBD_2021_DATA-855c6b27-1", 
                                 "IHME-GBD_2021_DATA-49677e16-1",
                                 "IHME-GBD_2021_DATA-6543986e-1", 
                                 "IHME-GBD_2021_DATA-c8c2f233-1",
                                 "IHME-GBD_2021_DATA-fedf4df8-1"),
    "Mental disorders" = c("IHME-GBD_2021_DATA-7e889f50-1",
                           "IHME-GBD_2021_DATA-9bcf5a9c-1",
                           "IHME-GBD_2021_DATA-733d6f1d-1",
                           "IHME-GBD_2021_DATA-773d5d49-1",
                           "IHME-GBD_2021_DATA-cc8dd461-1",
                           "IHME-GBD_2021_DATA-e6b878e8-1",
                           "IHME-GBD_2021_DATA-efa96b9d-1"),
    "Substance use disorders" = c("IHME-GBD_2021_DATA-3c475080-1",
                                  "IHME-GBD_2021_DATA-bf71612a-1",
                                  "IHME-GBD_2021_DATA-c3b9a773-1",
                                  "IHME-GBD_2021_DATA-def0a553-1",
                                  "IHME-GBD_2021_DATA-dfb0ffd2-1",
                                  "IHME-GBD_2021_DATA-e1890968-1",
                                  "IHME-GBD_2021_DATA-f0de7957-1"),
    "Diabetes and kidney diseases" = c("IHME-GBD_2021_DATA-4e038904-1",
                                       "IHME-GBD_2021_DATA-86e51847-1", 
                                       "IHME-GBD_2021_DATA-93ddb071-1",
                                       "IHME-GBD_2021_DATA-176ff4e4-1",
                                       "IHME-GBD_2021_DATA-598c99b4-1",
                                       "IHME-GBD_2021_DATA-b18d4bdf-1", 
                                       "IHME-GBD_2021_DATA-b94987d6-1"),
    "Skin and subcutaneous diseases" = c("IHME-GBD_2021_DATA-2ea9940f-1",
                                         "IHME-GBD_2021_DATA-2f416c57-1",
                                         "IHME-GBD_2021_DATA-6f2a84ba-1",
                                         "IHME-GBD_2021_DATA-9708e6f8-1",
                                         "IHME-GBD_2021_DATA-23375f3f-1",
                                         "IHME-GBD_2021_DATA-e29d941f-1",
                                         "IHME-GBD_2021_DATA-ef70347f-1"),
    "Sense organ diseases" = c("IHME-GBD_2021_DATA-397fe9b0-1", 
                               "IHME-GBD_2021_DATA-ccc561fc-1",
                               "IHME-GBD_2021_DATA-4aaa1bc9-1",
                               "IHME-GBD_2021_DATA-7e3893ce-1",
                               "IHME-GBD_2021_DATA-97ebd579-1",
                               "IHME-GBD_2021_DATA-547e86a9-1",
                               "IHME-GBD_2021_DATA-1165a657-1")
  ),
  output_name = "IHME-GBD_2021_CLEAN_prevalence",
  country = "United States"
)
```
