## Supplementary material for "Fisher-Rao distance and sex differences in disease prevalence trajectories": Code in R to reproduce the results: code_3.R

```
library(tidyverse)

fr_dist <- function(mu1, sigma1, mu2, sigma2, sqrt2 = 1.41421356237) {
  sqrt2 * acosh(1 + ((mu2 - mu1)^2 + 2 * (sigma2 - sigma1)^2) / (4 * sigma1 * sigma2))
}

load_region_data_aux <- function(file_path) {
  if (file.exists(file_path)) {
    readr::read_csv(file_path) |> janitor::clean_names()
  } else {
    stop("Data file not found: ", file_path) 
  }
}

load_region_data <- function(region) {
  load_region_data_aux(
    file_path = here::here("output", "code_1", paste0("IHME-GBD_2021_CLEAN_prevalence-", region, ".csv")) 
  )
}

compute_disease_distances <- function(df, dist_func, disease_name) {
  df |>
    filter(disease == disease_name) |>
    select(-disease) |>
    arrange(age) |>
    pivot_wider(
      id_cols = c(location, year, sex),
      names_from = age,
      values_from = c(mu, sigma),
      names_glue = "{.value}_{age}"
    ) |>
    mutate(
      total_dist = dist_func(mu_2, sigma_2, mu_7, sigma_7) +
        dist_func(mu_7, sigma_7, mu_12, sigma_12) +
        dist_func(mu_12, sigma_12, mu_17, sigma_17) +
        dist_func(mu_17, sigma_17, mu_22, sigma_22) +
        dist_func(mu_22, sigma_22, mu_27, sigma_27) +
        dist_func(mu_27, sigma_27, mu_32, sigma_32) +
        dist_func(mu_32, sigma_32, mu_37, sigma_37) +
        dist_func(mu_37, sigma_37, mu_42, sigma_42) +
        dist_func(mu_42, sigma_42, mu_47, sigma_47) +
        dist_func(mu_47, sigma_47, mu_52, sigma_52) +
        dist_func(mu_52, sigma_52, mu_57, sigma_57) +
        dist_func(mu_57, sigma_57, mu_62, sigma_62) +
        dist_func(mu_62, sigma_62, mu_67, sigma_67) +
        dist_func(mu_67, sigma_67, mu_72, sigma_72) +
        dist_func(mu_72, sigma_72, mu_77, sigma_77) +
        dist_func(mu_77, sigma_77, mu_82, sigma_82) +
        dist_func(mu_82, sigma_82, mu_87, sigma_87) +
        dist_func(mu_87, sigma_87, mu_92, sigma_92)
    ) |>
    select(location, year, sex, total_dist) |>
    mutate(is_male = as.integer(sex == "Male")) |>
    select(-sex) |>
    pivot_wider(
      id_cols = c(location, year),
      names_from = is_male,
      values_from = total_dist,
      names_glue = "{.value}_{is_male}"
    ) 
}

compute_regional_differences1 <- function(regions, dist_func, disease_list) {
  map_dfr(regions, ~ {
    df <- load_region_data(.x)
    map_dfr(disease_list, ~ compute_disease_distances(df, dist_func, .x) |>
              mutate(d = total_dist_1 - total_dist_0) |>
              select(-total_dist_1, -total_dist_0) |>
              group_by(year) |>
              summarise(
                g = (mean(d > 0) - 0.5) |> round(digits = 4),
                mu = mean(d) |> round(digits = 4),
                sigma = sd(d) |> round(digits = 4),
                skewness = e1071::skewness(d) |> round(digits = 4),
                kurtosis = e1071::kurtosis(d) |> round(digits = 4),
                shapiro_p = shapiro.test(d) |> getElement("p.value") |> round(digits = 4),
                t_p = t.test(d, alternative = "two.sided") |> getElement("p.value") |>  round(digits = 4)
              ) |>
              mutate(disease = .x)) |>
      mutate(region = .x)
  })
}

DISEASE_LIST <- c("Neoplasms", "Cardiovascular diseases", "Chronic respiratory diseases", "Digestive diseases",
                 "Neurological disorders", "Mental disorders", "Substance use disorders", "Diabetes and kidney diseases",
                 "Skin and subcutaneous diseases", "Sense organ diseases")

df <- compute_regional_differences1(
      regions = c("United States", "Western Europe", "Japan"), 
      dist_func = fr_dist,
      disease_list = DISEASE_LIST
    )

sapply(DISEASE_LIST, function(disease0) {
  df |> 
    filter(year == 1990, disease == disease0) |>
    select(-year, -disease) |> 
    select(region, g, mu, sigma, skewness, kurtosis, shapiro_p, t_p) |>
    xtable::xtable() |>
    print(include.rownames = FALSE,
          file = fs::path(here::here("output","code_3",disease0), ext = "tex"))
})
```
