## Supplementary material for "Fisher-Rao distance and sex differences in disease prevalence trajectories": Code in R to reproduce the results: code_4.R

```
library(tidyverse)

Fisher_Rao <- function(mu1, sigma1, mu2, sigma2, sqrt2 = 1.41421356237) {
  sqrt2 * acosh(1 + ((mu2 - mu1)^2 + 2 * (sigma2 - sigma1)^2) / (4 * sigma1 * sigma2))
}

create_df <- function(age, d_male, d_female) {
  data.frame(
    age = age,
    u_male = cumsum(d_male),
    u_female = cumsum(d_female)
  )
}

process_sex_aux <- function(df_male, df_female, f) {
  create_df(
    age = tail(df_male, -1)$age,
    d_male = sapply(2:nrow(df_male), function(i)
      f(
        mu1 = df_male$mu[i-1], sigma1 = df_male$sigma[i-1],
        mu2 = df_male$mu[i], sigma2 = df_male$sigma[i]
      )
    ),
    d_female = sapply(2:nrow(df_female), function(i)
      f(
        mu1 = df_female$mu[i-1], sigma1 = df_female$sigma[i-1],
        mu2 = df_female$mu[i], sigma2 = df_female$sigma[i]
      )
    )
  )
}

process_sex <- function(df_disease, f, loc, yr) {
  process_sex_aux(
    df_male = df_disease |>
      filter(location == loc, year == yr, sex == "Male") |>
      select(age, mu, sigma) |>
      arrange(age), 
    df_female = df_disease |>
      filter(location == loc, year == yr, sex == "Female") |>
      select(age, mu, sigma) |>
      arrange(age), 
    f = f
    )
}

create_graphics <- function(plot_df, disease0, loc, yr) {
  plot_df |>
  ggplot(aes(x = age)) +
    geom_line(aes(y = u_male, color = "Male", linetype = "Male"), size = 1.5) +
    geom_line(aes(y = u_female, color = "Female", linetype = "Female"), size = 1.5) +
    labs(
      title = paste(disease0, loc, yr, sep = " - "),
      x = "Age (±2 years)",
      y = "Cumulative Fisher-Rao Distance",
      color = "Sex",
      linetype = "Sex",
      caption = "Source: GBD-2021 (IHME)"
    ) +
    scale_color_manual(values = c("Male" = "black", "Female" = "gray")) +
    scale_linetype_manual(values = c("Male" = "solid", "Female" = "solid")) +
    theme_bw() +
    theme(legend.position = "bottom") +
    scale_x_continuous(breaks = sort(unique(plot_df$age)))
}

process <- function(df_disease, disease0, loc, yr) {
  ggsave(
    filename = here::here("output", "code_4", paste(
      gsub(" ", "_", disease0),
      gsub(" ", "_", loc),
      yr,
      sep = "_"
    ) |> paste0(".png")),
    plot = create_graphics(process_sex(
      df_disease = df_disease, 
      f = Fisher_Rao,
      loc = loc,
      yr = yr
    ) |> mutate(age = age - 5),
    disease0 = disease0,
    loc = loc,
    yr = yr
    ),
    width = 8,
    height = 6,
    dpi = 300
  )
}

main_aux <- function(df_data, diseases, loc, yr) {
  sapply(diseases, function(disease0) {
    process(
      df_disease = df_data |> 
        filter(disease == disease0) |> 
        select(-disease),
      disease0 = disease0,
      loc = loc, 
      yr = yr
    )
  })
}

main <- function(seed, diseases, loc, yr, filename_in) {
  withr::with_seed(seed,
      main_aux(
        df_data = readr::read_csv(fs::path(here::here("output", "code_1",
          filename_in), ext = "csv")),
        diseases = diseases,
        loc = loc,
        yr = yr
      )
  )
}

main(
  seed = 123,
  diseases = c("Chronic respiratory diseases"),
  loc = "California",
  yr = 1990,
  filename_in = "IHME-GBD_2021_CLEAN_prevalence-United States"
)
```
