## Supplementary material for "Fisher-Rao distance and sex differences in disease prevalence trajectories": Code in R to reproduce the results: code_5.R

```
library(tidyverse)

create_graphics <- function(plot_df, disease0, loc, yr, sex0) {
  plot_df <- plot_df |>
    mutate(
      mu_next = lead(mu),
      sigma_next = lead(sigma),
      mid_mu = (mu + mu_next) / 2,
      mid_sigma = (sigma + sigma_next) / 2
    )
  
  ggplot(plot_df, aes(x = mu, y = sigma, label = age)) +
    geom_point(color = "black", size = 2) +
    geom_segment(
      aes(x = mu, y = sigma, xend = mid_mu, yend = mid_sigma),
      arrow = arrow(length = unit(0.2, "cm")),
      color = "darkgray",
      size = 0.5
    ) +
    geom_segment(
      aes(x = mid_mu, y = mid_sigma, xend = mu_next, yend = sigma_next),
      color = "darkgray",
      size = 0.5
    ) +
    labs(
      title = paste(disease0, loc, yr, sex0, sep = " - "),
      x = "μ",
      y = "σ",
      caption = "Source: GBD-2021 (IHME)"
    ) +
    xlim(-1.4, 0) + # change according to the disease and location
    ylim(0.02, 0.13) + # change according to the disease and location
    theme_bw()
}

process <- function(df_disease, disease0, loc, yr) {
  sapply(c("Male", "Female"), function(sex0) {
    ggsave(
      filename = here::here("output","code_5", paste(
        gsub(" ", "_", disease0),
        gsub(" ", "_", loc),
        yr,
        sex0,
        sep = "_"
      ) |> paste0(".png")),
      plot = create_graphics(
        plot_df = df_disease |>
          filter(sex == sex0) |>
          select(age, mu, sigma) |>
          arrange(age),
        disease0 = disease0,
        loc = loc,
        yr = yr,
        sex = sex0
      ),
      width = 8,
      height = 6,
      dpi = 300
    )
  })
}

main_aux <- function(df_data, diseases, loc, yr) {
  sapply(diseases, function(disease0) {
    process(
      df_disease = df_data |>
        filter(disease == disease0, location == loc, year == yr) |>
        select(-disease),
      disease0 = disease0,
      loc = loc,
      yr = yr
    )
  })
}

main <- function(seed, diseases, loc, yr, filename_in) {
  withr::with_seed(seed,
                   main_aux(
                     df_data = readr::read_csv(
                       fs::path(here::here("output", "code_1",
                                                filename_in), ext = "csv")),
                     diseases = diseases,
                     loc = loc,
                     yr = yr
                   )
  )
}

main(
  seed = 123,
  diseases = c("Chronic respiratory diseases"),
  loc = "California",
  yr = 1990,
  filename_in = "IHME-GBD_2021_CLEAN_prevalence-United States"
)
```
